# Robustly Quantifying Uncertainty in International Avian Influenza A(H5N1) Infection Fatality Ratios

**DOI:** 10.64898/2026.04.22.26351373

**Authors:** Leonardo Gada, Mwandida Kamba Afuleni, Michael Noble, Thomas House, Thomas Finnie

## Abstract

Knowing the mortality rates associated with infection by a pathogen is essential for effective preparedness and response. Here, harnessing the flexibility of a Bayesian approach, we produce an estimate of the Infection Fatality Ratio (IFR) for A(H5N1) conditional on explicit assumptions, and quantify the uncertainty thereof. We also apply the method to first-wave COVID-19 data up to March 2020, demonstrating the estimates that could be obtained were the model available then.

Our analysis uses World Development Indicators (WDI) from the World Bank, the A(H5N1) WHO confirmed cases and deaths tracker by country (2003-2024), and COVID-19 cases and deaths data from John Hopkins University (January and February 2020). Since infectious disease dynamics are typically influenced by local socio-economic factors rather than political borders, individual countries are placed within clusters of countries sharing similar WDIs relevant to respiratory viral diseases, with clusters derived by performing Hierarchical Clustering. To estimate the IFR, we fit a Negative Binomial Bayesian Hierarchical Model for A(H5N1) and COVID-19 separately. We explicitly modelled key unobserved parameters with informative priors from expert opinion and literature.

By modelling underreporting, our analysis suggests lower fatality (15.3%) compared to WHO’s Case Fatality Ratio estimate (54%) on lab-confirmed cases. However, credible intervals are wide ([0.5%, 64.2%] 95% CrI). Therefore, good preparedness for a potential A(H5N1) pandemic implies adopting scenario planning under our central estimate, as well as for IFRs as high as 70%. Our approach also returns a COVID-19 IFR estimate of 2.8% with [2.5%, 3.1%] 95% CrI which is consistent with literature.

**Key Messages:**

1. We adopted a disease-agnostic and adaptable Bayesian model, embedding scientific knowledge on A(H5N1) in the priors informed by published literature, to estimate the Infection Fatality Ratio (IFR) of avian influenza A(H5N1).
2. Accounting for underreporting of cases and deaths, we estimate the IFR of avian influenza A(H5N1) at 15.3%, albeit with wide uncertainty ([0.5%, 64.2%] 95% Credible Intervals).
3. Due to the uncertainty in the estimate, good preparedness for a potential A(H5N1) pandemic implies adopting scenario planning under our central estimate, as well as for IFRs as high as 70%.

## Introduction

Avian influenza A(H5N1) is an enveloped RNA virus of the *Orthomyxoviridae* family which is highly pathogenic in birds, particularly poultry, and has been detected in several mammalian species including relatively recently dairy cattle [1]. Certain strains have also spilled over into humans causing severe respiratory infection, with the first recognised human outbreak reported in Hong Kong in 1997, resulting in 18 confirmed cases and six deaths [2]. H5N1 has since become endemic in poultry populations across Asia, the Middle East and Africa, resulting in repeated spillovers to humans [2, 3].

Recent outbreaks have increased global concern regarding the pandemic potential of H5N1. Between January 1 and August 4, 2025, 26 human infections were reported worldwide, including 11 fatalities (8 in Cambodia; 2 in India; and 1 in Mexico) with most cases associated with direct or likely contact with infected poultry or wild birds [4]. In the United States, widespread outbreaks of H5N1 viruses have been reported among poultry, dairy cattle, and other animals, with 70 human infections documented during 2024 and early 2025 [4]. Cumulatively, the World Health Organization (WHO) has documented 976 laboratory-confirmed human H5N1 infections and 470 deaths globally between 2003 and May 2025, corresponding to an overall raw case fatality rate (CFR) of 48% [5].

More accurately quantifying the mortality burden associated with H5N1 viruses is challenging, as WHO surveillance systems are restricted to report only laboratory-confirmed cases, leading to substantial underestimation of both the incidence and mortality associated with H5N1 [6]. This underreporting limits the ability to produce reliable estimations of the infection fatality ratio (IFR), which is defined as the risk of death among all infections, including those not detected as cases, for example due to being asymptomatic [7]. Serological studies have sought to address this gap and refine IFR estimates by detecting past exposure through antibody prevalence, however, these studies have produced conflicting results. Some studies report very low H5N1 antibody prevalence, suggesting that subclinical infection in humans are rare [8], whereas others, such as a recent serological study among U.S. dairy workers, report higher seropositivity, indicating more frequent asymptomatic infections in certain settings [9]. A recent paper by Drake [10] discussed different hypotheses for the patterns of observed mortality associated with H5N1, highlighting the persistent uncertainty on the extent of asymptomatic cases, and hence of the suitability of the CFR as opposed to IFR as a measure of fatality.

H5N1 viruses lack the ability to efficiently bind to receptors that are predominant in the human upper respiratory tract, thus limiting their capacity for efficient human-to-human transmission and keeping the overall pandemic risk low [11]. However, genetic data have shown that when mammals, including humans, are infected, the virus can undergo intra-host evolution resulting in changes that enable more efficient replication in the lower respiratory tract [12]. For example, the PB2-E627K marker was detected in human cases in Cambodia, Vietnam, and the United States, a mutation strongly associated with mammalian adaptation during infection [9, 12, 13]. These developments have led to heightened surveillance and preparedness measures by public health agencies globally, including in the United Kingdom, where risk assessments have emphasised the potential for H5N1 to evolve into a virus capable of sustained human-to-human transmission [6].

This study aims to estimate the IFR for A(H5N1) accounting for underreporting of cases and deaths, as well as quantifying the uncertainty of said estimate. Due to the limits and sparsity in the current knowledge base, we build a two-level Bayesian Hierarchical Model which allows us to incorporate sources of information such as expert opinion and the results of other studies explicitly in the form of parameter priors.

## Methodology

Multiple datasets underpin our analysis. For A(H5N1) we rely on the WHO tracker of cumulative yearly cases and deaths by country from January 2003 to March 2024 [14], and for COVID-19 on the John Hopkins University’s daily timeseries of cases and deaths by country (very early phase of the pandemic, January and February 2020) [15]. Although this study focuses on A(H5N1), we use COVID-19 data as a comparator to validate the modelling approach since fatality rates are much less uncertain. While much is still unknown about Influenza A(H5N1), our knowledge of COVID-19 is vast and provides us with a suitable benchmark.

Since infectious disease epidemiology is influenced by socio-demographic factors, individual countries are placed within clusters of countries sharing similar characteristics. The clustering is based on World Development Indicators (WDI) from the World Bank Database relevant to respiratory viral diseases. The full procedure is described in Afuleni, Gada et al. [16]. The cluster map and cluster memberships are available to view in the Appendix.

To account for population size in different clusters, we use yearly population counts from the World Bank Database. For COVID-19 these are the 2019 population counts.

The model description in this section follows recommendations outlined in Depaoli & van de Schoot [17].

### Model Structure

In our framework, we anchor the model to reality via Negative Binomial log likelihood functions for cases (*C*) and deaths (*D*) separately:

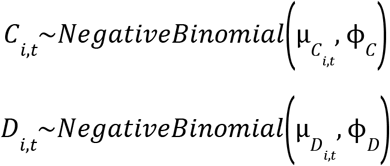

Where *i* ∈{*Cluster*1, …, *ClusterN*} and *t* ∈{*Year*1, …, *YearN*} and we use the parameterisation of the Negative Binomial in terms of a mean and overdispersal in *Stan*’s *NegBinomial2()* function. The means are derived parameters from the true number of cases and deaths, multiplied by the probability of reporting:

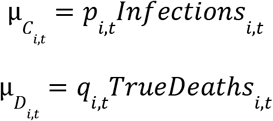

The true number of cases (infections) and deaths are derived sequentially as follows, highlighting the role played by our object of interest, the true IFR:

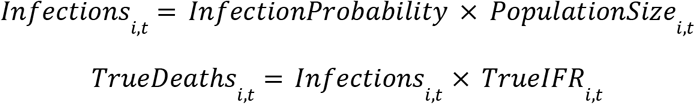

The probabilities of reporting (*p, q*) are modelled using a logit link function, and encode the hierarchical structure with random effects for cluster (betas) and year (gammas):

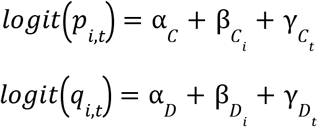

### Priors

Table 1 outlines the full list of parameters and priors used in the two models.

**Table 1.** Full list of parameters and priors. A(H5N1) and COVID-19 model. Where N is the number of cluster-year combinations in our dataset for A(H5N1) and the number of clusters for COVID-19.

| Parameter Name | A(H5N1) Model Prior | COVID-19 Model Prior | Description |
| --- | --- | --- | --- |
| Infection Probability | Beta (0.5, 100) | Beta (0.5, 100) | Probability of infection |
| True IFR | Beta (1.25, 2) | Beta (0.96, 47) | Real IFR, our target |
| Alpha cases, $\alpha_c$ | Normal (0, 100) | Normal (0, 100) | Baseline reporting rate for cases |
| Alpha deaths, $\alpha_D$ | Normal (0, 100) | Normal (0, 100) | Baseline reporting rate for deaths |
| Phi cases, $\Phi_c$ | Gamma (1, 1) | Gamma (1, 1) | Dispersion for distribution of cases |
| Phi deaths, $\Phi_D$ | Gamma (1, 1) | Gamma (1, 1) | Dispersion for distribution of deaths |
| Beta cases standardised | Normal (0, 1) | Normal (0, 1) | Cluster random effect for cases |
| Beta deaths standardised | Normal (0, 1) | Normal (0, 1) | Cluster random effect for deaths |
| Beta sigma cases | Half Cauchy (0, 2.5) | Half Cauchy (0, 2.5) | Standard deviation for Beta cases |
| Beta sigma deaths | Half Cauchy (0, 2.5) | Half Cauchy (0, 2.5) | Standard deviation for Beta deaths |
| Beta cases, $\beta_c$ | Flat | Flat | Non-standardised Beta cases |
| Beta deaths, $\beta_D$ | Flat | Flat | Non-standardised Beta deaths |
| Gamma cases standardised | Normal (0, 1) | N/A | Year random effect for cases |
| Gamma deaths standardised | Normal (0, 1) | N/A | Year random effect for deaths |
| Gamma tau cases | Half Cauchy (0, 2.5) | N/A | Standard deviation for Gamma cases |
| Gamma tau deaths | Half Cauchy (0, 2.5) | N/A | Standard deviation for Gamma deaths |
| Gamma cases, $\gamma_c$ | Flat | N/A | Non-standardised Gamma cases |
| Gamma deaths, $\gamma_D$ | Flat | N/A | Non-standardised Gamma deaths |
| Infections | Gamma (1.85, 0.01) | Gamma (324, 0.003) | Total number of cases per cluster-year |
| Mu cases, $\mu_c$ | Gamma (0.54, 0.02) | Gamma (0.91, 4.27e-5) | Negative Binomial parameter for cases |
| Mu deaths, $\mu_D$ | Gamma (0.47, 0.04) | Gamma (1.10, 1.50e-3) | Negative Binomial parameter for deaths |
| Case reporting probability, $p$ | Beta (1.25, 3) | Beta (2, 2) | Reporting probability for cases |
| Death reporting probability, $q$ | Beta (3, 1.50) | Beta (2, 2) | Reporting probability for deaths |
| True deaths | Gamma (1, 0.06) | Gamma (3600, 1.20) | Total number of deaths per cluster-year |

Our approach is explicitly designed to use a Bayesian approach to integrate existing knowledge to mitigate the small volumes of data available, meaning we did not use uniform, improper or otherwise uninformative priors where possible.Instead, we have taken priors from data, as well as experts and the scientific literature. The two sections below describe the sources and rationale for the informative priors.

### H5N1 Model

The infection probability was given an uninformative prior based on seasonal flu by expert elicitation, while the true IFR was informed by Ward et al. [18]. Infections, true deaths, 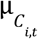 and 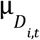 were assigned an appropriate distribution, the parameters of which we derived with the method of moments. The μ distributions were centred on the mean of observed cases and observed deaths calculated from the dataset, while their variance was set at 1000 and 300 respectively after observing the distribution of the data. We followed a similar reasoning for Infections and true deaths. Since we assume underreporting (greater for cases than deaths), infections were centred on the maximum number of cases from the data with standard deviation of 100, while true deaths were centred on the mean number of deaths + 5 and had a standard deviation of 17. We used literature reviewing surveillance systems during the first wave of COVID-19 [19][20][21] to inform the probability of reporting of cases, while we assumed deaths to be more evident than infections and therefore more accurately reported, albeit not perfectly. The global intercepts (α_*C*_ and α_*D*_) and the dispersion parameters (ϕ_*C*_ and ϕ_*D*_) received an uninformative prior compatible with their support. The random effects parameters (β_*C*_, β_*D*_, γ_*C*_, γ_*D*_) were specified with non-centred parametrisation [22] for ease of computation and given a flat prior upon definition. This entails specifying, for each parameter, two additional subservient “Mean” and two “Standard deviation” parameters that are also listed in Table 1, showing priors compatible with the reparametrisation procedure and their support.

### COVID-19 Model

The COVID-19 model is specified as if we were in March 2020. It does not incorporate any knowledge that would have been available only after March 2020. The infection probability is informed by seasonal flu, while the true IFR is based on the SPI-M-O Consensus Statement of February 2020 [23]. Infections, true deaths, 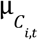 and 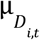 were assigned an appropriate distribution, the parameters of which we derived with the method of moments. The μ distributions were centred on the mean of observed cases and observed deaths from the John Hopkins Dashboard with standard deviation set at 10,000 and 700 respectively. We again apply the method of moments to set mean infections at approximately 10,000 (standard deviation 6000) and true deaths at 3000 (standard deviation 50). The probability of reporting of cases and deaths are given an uninformative Beta prior, since we have no plausible benchmark. Global intercepts and dispersion parameters are treated as above. We only model a random effect with non-centred parametrisation for clusters as all observations are from 2020 (Table 1).

### Computation and Diagnostics

We carried out the analysis in R 4.4.2 “Pile of Leaves” and Stan via the *CmdStanR* package [24]. Both models are fitted with the same specifications using the No-U-Turn-Sampler (NUTS) algorithm, the default Markov Chain Monte Carlo (MCMC) method in Stan. We run four parallel chains with 5000 warm-up steps and 10,000 iterations each. The adaptive delta was set at 0.99 and the step size at 0.01. We stored CmdStanR output for the number of divergences, tree depth hits and the estimated Bayesian fraction of missing information (EBFMI). We then used this to look at the effective sample size (ESS) and R-hat for each parameter to rule out any obvious fitting issues, as well as autocorrelation plots and the NUTS energy plot. Chain mixing is assessed via trace plots with settings as specified and with twice the number of iterations to rule out local convergence. Global convergence is additionally assessed by inspecting the Geweke-Brooks plots via the *coda* package [25]. We also inspect the posterior histograms to verify the iterations offered an adequate representation of the posterior. We perform posterior predictive checks (PPC) using the *bayesplot* package [26][27], and prior sensitivity checks (PSC) with the *priorsense* package [28], which implements power scaling.

## Results

The overall IFR for A(H5N1) is estimated, under our modelling assumptions, at 15.3% with 95% CrI as [0.51%, 64.2%]. The model also calculates a distinct IFR posterior distribution for each of the 39 cluster-year combinations (Figure 1 and 2). The cluster-year dictionary is in the Appendix (Tables A1 and A2).

**Figure 1.**
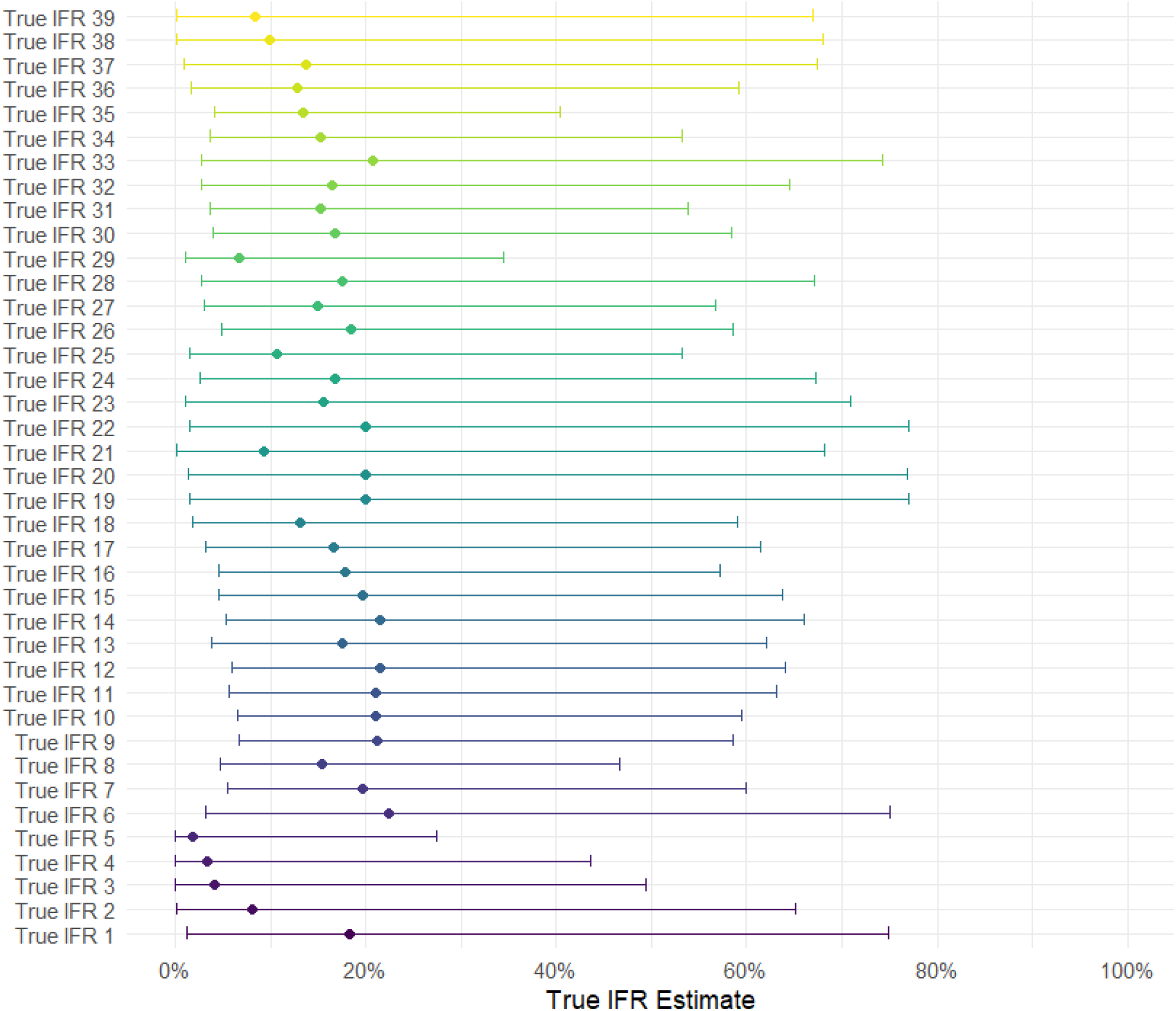
Estimates of each cluster-year IFR. The dot is the median, while the thin line indicates the 95% CrI.

**Figure 2.**
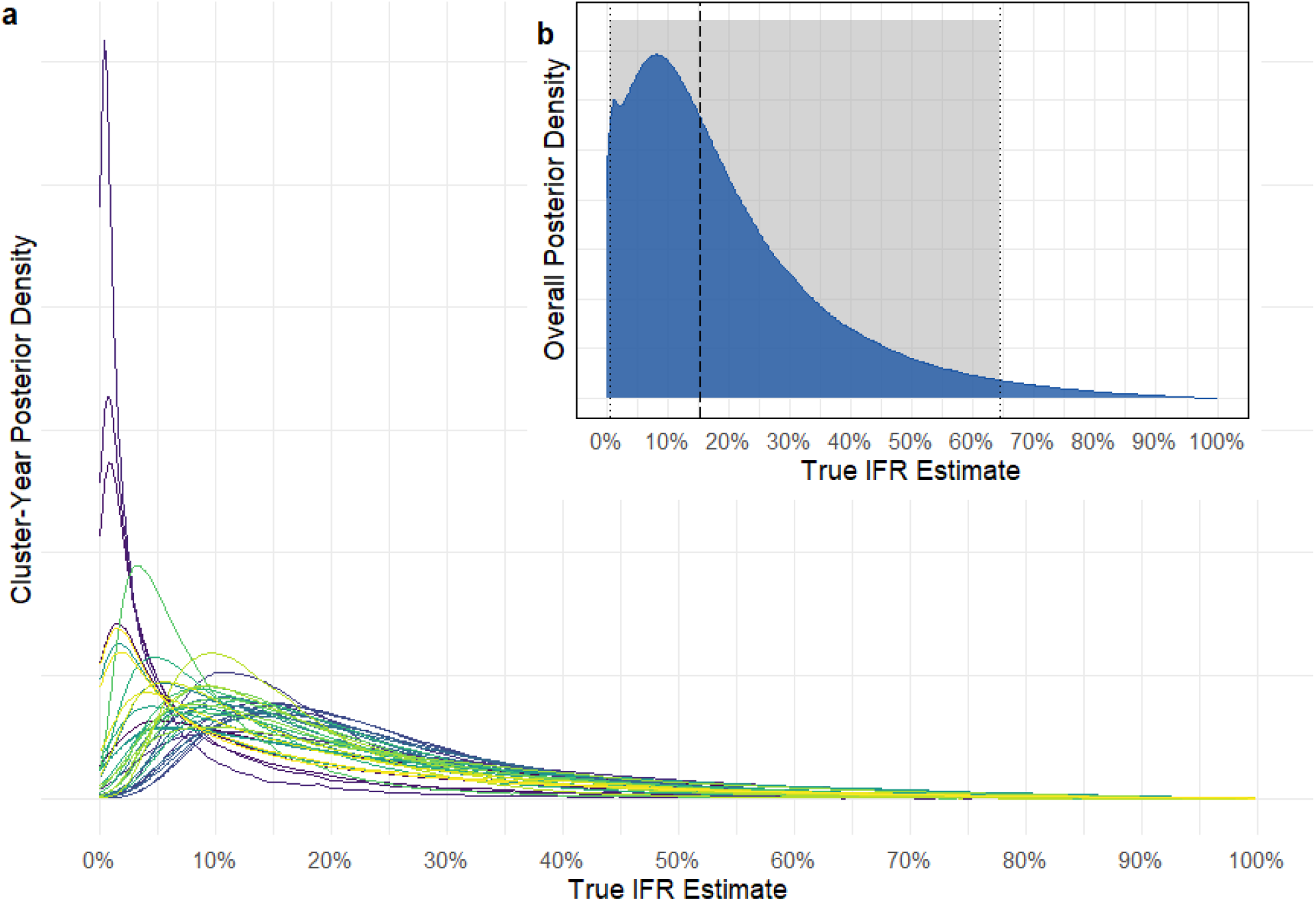
Density plots for cluster-year A(H5N1) IFRs. Each line at (a) is a cluster-year pair, while (b) shows the overall posterior density of the IFR estimate with shaded 95% CrI. The dashed line is the median value.

Diagnostics indicate no issues with convergence nor autocorrelation, and the EBFMI is stable at around 0.80. The Geweke-Brooks plots (in Supplementary Material) show that all parameters in all chain remain within the acceptable region, or wander near the borders. There is one divergent transition (0.0% of total transitions) which we have investigated but found no reasons for apart from random occurrence. The PPCs show good fit for both cases and deaths replicates (Figure 3).

**Figure 3.**
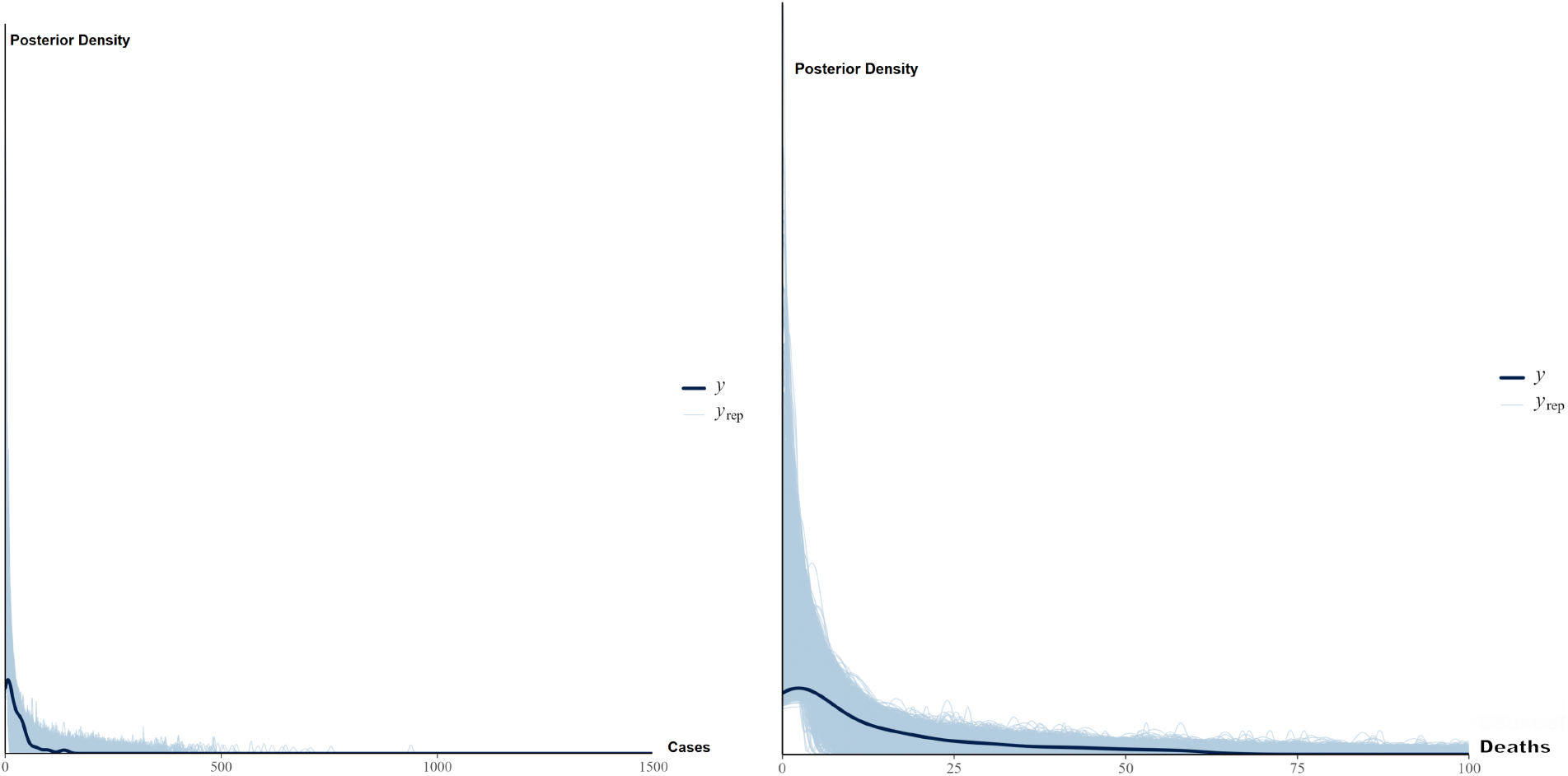
Shading indicates the density distribution of replicates of cases (left) and deaths (right) from the H5N1 model with overlaid observed values (solid blue line).

**Figure 4.**
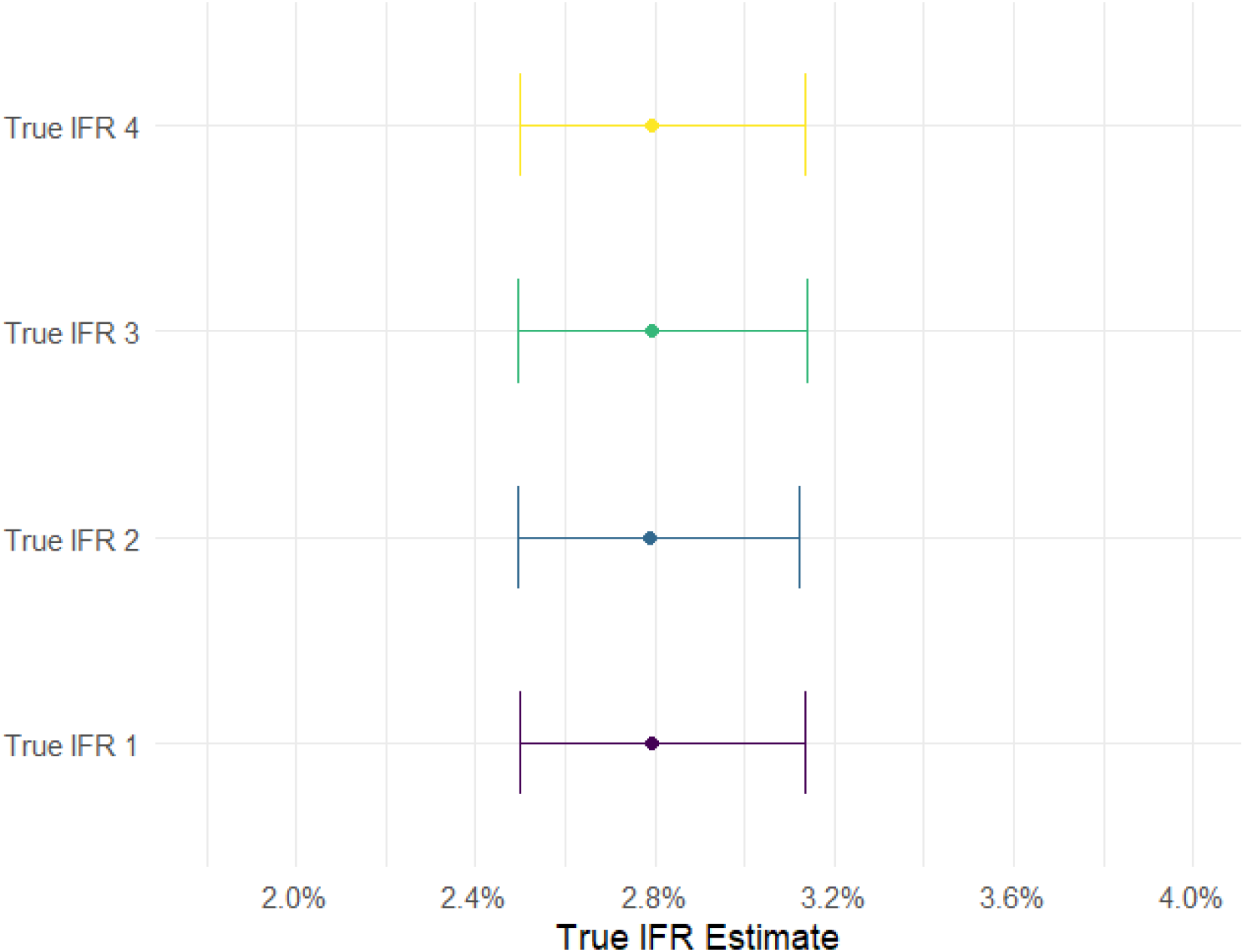
Estimates of each cluster’s IFR. The dot is the median, while the thin line indicates the 95% CrI.

**Figure 5.**
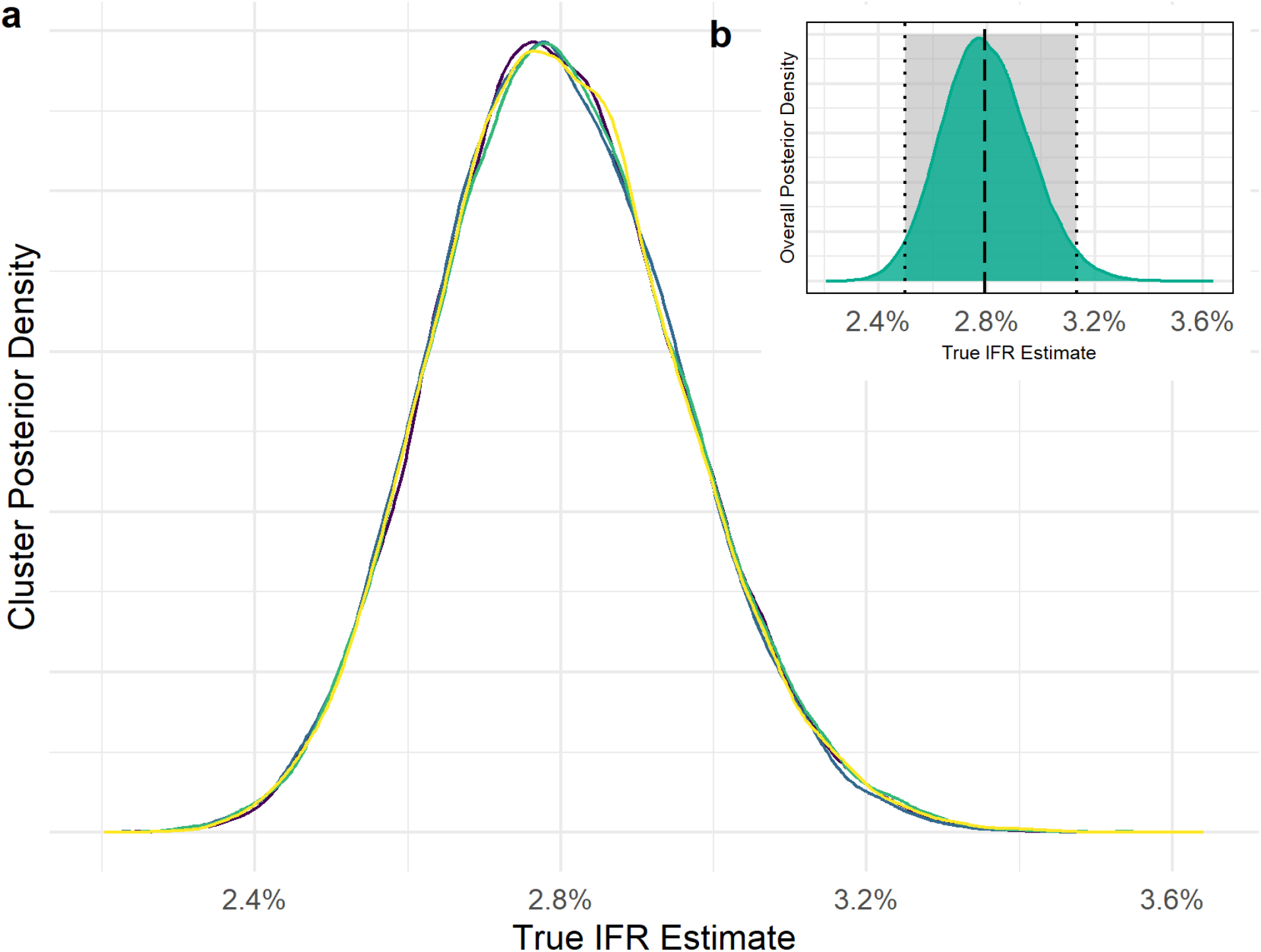
Density plots for cluster COVID-19 IFRs. Each line at (a) is a cluster, while (b) shows the overall posterior density of the IFR estimate with shaded 95% CrI. The dashed line is the median value.

**Figure 6.**
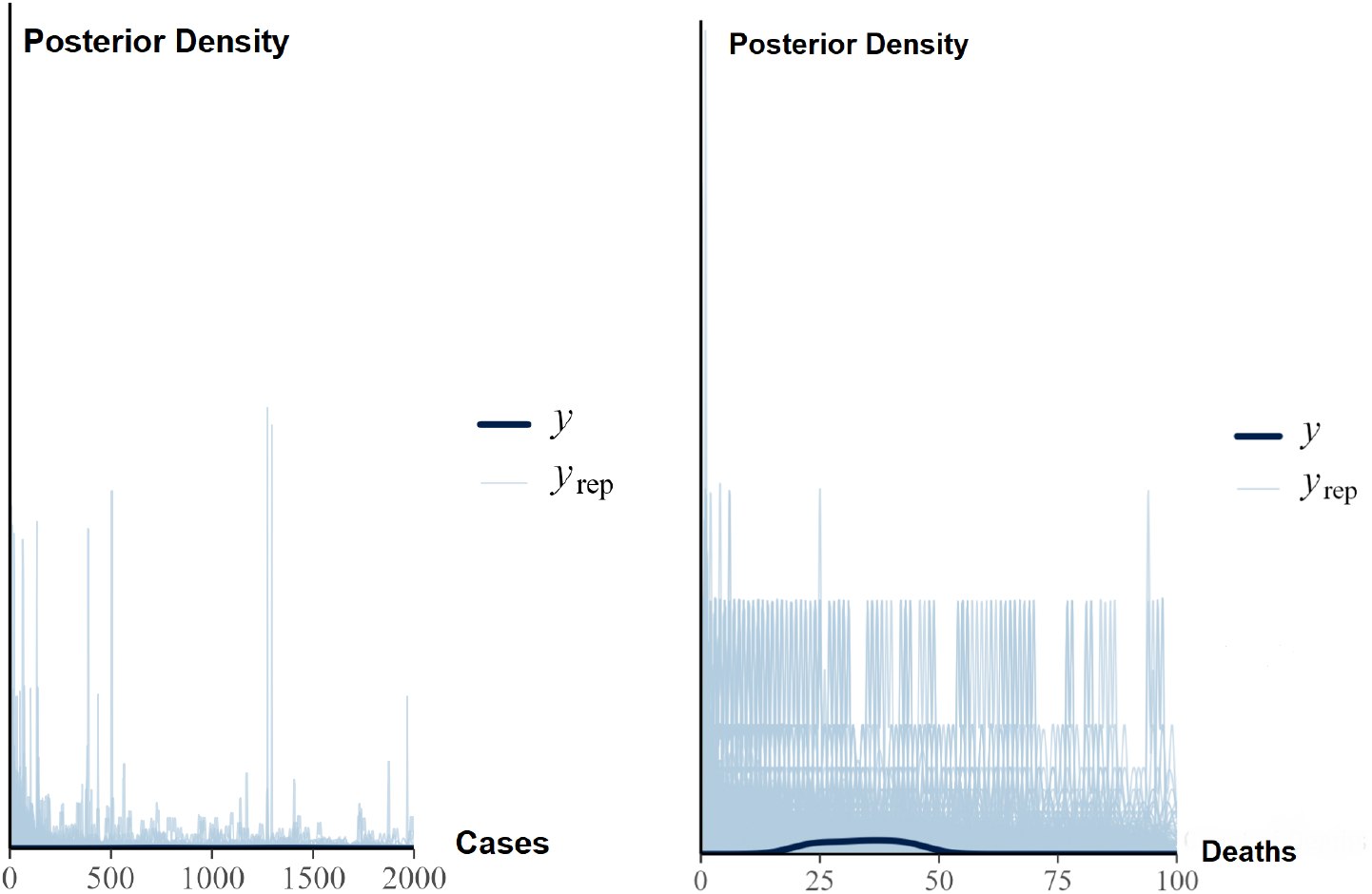
Shading indicates the density distribution replicates of cases (left) and deaths (right) from the COVID-19 model with overlaid observed values (solid blue line).

The PSC flag some potential prior-data conflicts. However, upon inspection of the *priorsense* plots for power-scaled priors and likelihood (in Supplementary Material), we do not see evident sensitivities. The developers suggest treating the threshold triggering diagnostic messages as a context-dependent guideline only, in this case indicating the default threshold might not be suitable [29].

The comparator COVID-19 model ran with no issues of convergence nor autocorrelation (plots in Supplementary Material). The EBFMI was stable around 0.75 for all chains and there were no divergent transitions. The overall COVID-19 IFR estimate was 2.8% with 95% CrI [2.5%, 3.1%].

The PSC found no potential prior sensitivity issues via power scaling. The PPC highlighted this model struggles more with predictive power, as shown by the blips in the plots with superimposed replicates and observed values. For cases, the model’s overshoots are significant enough to make the observed values unreadable given their flattening against the x axis. This is likely due to the COVID-19 dataset containing only four observations (one row per cluster) since we removed the time dimension [30]. As we use the COVID-19 model as a comparison only, we are not concerned by the PPC result given it’s most likely driven by the low number of observations in the dataset.

## Discussion

Our study demonstrates a flexible modelling framework ideal to reckon with the inherent limitations of the present knowledge base on A(H5N1). Differently from the frequentist paradigm, Bayesian modelling is more viable with small datasets and allows for direct incorporation of knowledge from literature to partially counterbalance this. The lack of scientific consensus on the likelihood and incidence of mild and asymptomatic A(H5N1) cases from antibody-based studies steered us away from the use of serological information to correct for underreporting. Our search of literature on goodness of surveillance based on retrospective post-covid studies provided us with a reasonable, readily available baseline for reporting of cases and deaths in the countries of interest. The model fit and convergence passed standard checks. The resulting IFR and Credible Interval provide a revised estimate of A(H5N1) fatality that incorporates a transparent quantification of the uncertainty. We feel this is an honest assessment of our current knowledge of this pathogen.

As with any scientific study, there are limitations. A disease’s fatality varies greatly by age group, however, due to the WHO tracker being aggregated by country and year, we could not include any individual-level demographics in the model. Line list data would be better to examine the variation of fatality by attributes such as age, gender and comorbidities. Our data also does not include the US outbreak of 2024 due to the timing of the study and data upload on the WHO website. Since the US outbreak was mainly expressed via conjunctivitis symptoms rather than respiratory, we do not deem this to be an issue, however we could not verify what other past cases in the tracker might have not been respiratory in nature.

Caution should be used in extending the interpretation of parameters from the model to the whole cluster they are part of, and to individual countries within them, with particular attention to those countries for which no data were in the dataset. For example, Turkey is in the same cluster as Mexico, but while Turkey had observed cases and contributed to the model, Mexico had no cases and therefore didn’t contribute to the generation of the estimates. This is because the clustering was performed on all the countries in the World Development Indicators dataset, as outlined in a separate paper [16], and then simply used for grouping in this analysis. As previously discussed, the indicators used as foundation for the clustering algorithm were chosen based on their relevance for a respiratory disease. Hence, the clustering should not be uncritically applied to parameter estimation for non-respiratory diseases. The reader can view the full table of IFR estimate ID, clusters, and cluster memberships in the Appendix.

We also recognise some of our assumptions can be challenged, such as the use of retrospective literature on goodness of surveillance during the COVID-19 pandemic as a proxy for probability of reporting cases and deaths for A(H5N1). Vital records completeness estimates could be an alternative, such as those from the Global Burden of Disease (GBD) study [31]. However, we considered our proxy to be closer to the reality of public health surveillance for avian influenza, especially for respiratory symptoms. Practically, we would suggest that in planning, alternative priors could be asserted - for example depending on which of the three hypotheses for H5N1 mortality patterns identified by Drake [10] is assumed to hold - with our modelling framework then allowing for principled propagation of uncertainty under those assumptions.

This model is a starting point and can be easily rerun as more data becomes available. Future improvements could include the introduction of subtyping information to account for differences in A(H5N1) variants between outbreaks, the introduction of a pooled measure of records completeness by country to derive the overall cluster’s measure, and the development of a module to allow modelling of policy interventions. It will be crucial to remain mindful of the risks of overparametrisation due to the paucity of data. The Bayesian approach is flexible, but with no solid anchor to reality via the data all that remains are the assumptions encoded in the priors.

## Conclusion

Our study presents an honest assessment of current knowledge concerning overall IFRs for A(H5N1). Although our model suggests lower fatality (15.3%) compared to WHO’s CFR estimate (54%) on lab-confirmed cases, credible intervals are wide ([0.5%, 64.2%] 95% CrI). This reflects the many unknowns on avian influenza, indicating that pandemic preparedness should not be built based on a single point estimate. Doing so would not only misrepresent reality, but also build a harmful false sense of security in decision makers. Instead, good preparedness for a potential A(H5N1) pandemic implies adopting scenario planning under our central estimate, as well as for IFRs as high as 70%. The analytical methodology we deployed here is disease-agnostic, and as such it could be easily repurposed for other pathogens and similar parameter estimation.

## Supporting information

Supplementary Diagnostic Plots

## Data Availability

The data used in this analysis are publicly available online as well as the scripts used to produce the results

https://www.who.int/publications/m/item/cumulative-number-of-confirmed-human-cases-for-avian-influenza-a(h5n1)-reported-to-who--2003-2025--20-january-2025]

https://github.com/CSSEGISandData/COVID-19/tree/master/csse_covid_19_data/csse_covid_19_time_series

https://github.com/ukhsa-collaboration/ModellingInfectionFatalityRatiosH5N1/tree/main

## Funding statement

MKA is supported by the Schlumberger Foundation–Faculty for the Future. TH is supported by the Wellcome Trust (Ref: 227438/Z/23/Z) and Medical Research Council (Ref: UKRI483).

LG, MN, TF are employed by UKHSA. The research leading to these results received UK Government grant-in-aid funding to UKHSA. The views expressed in this publication are those of the authors and not necessarily those of UKHSA or Department for Health and Social Care.

The funders had no role in study design, data collection and analysis, decision to publish, or preparation of the manuscript.

## Acknowledgements

We would like to thank Rabia A. Khan, Ailbe Nolan for contributing epidemiological comments and insights to our work and Martyn Fyles for advice on Stan code.

## APPENDIX

**Figure A1.**
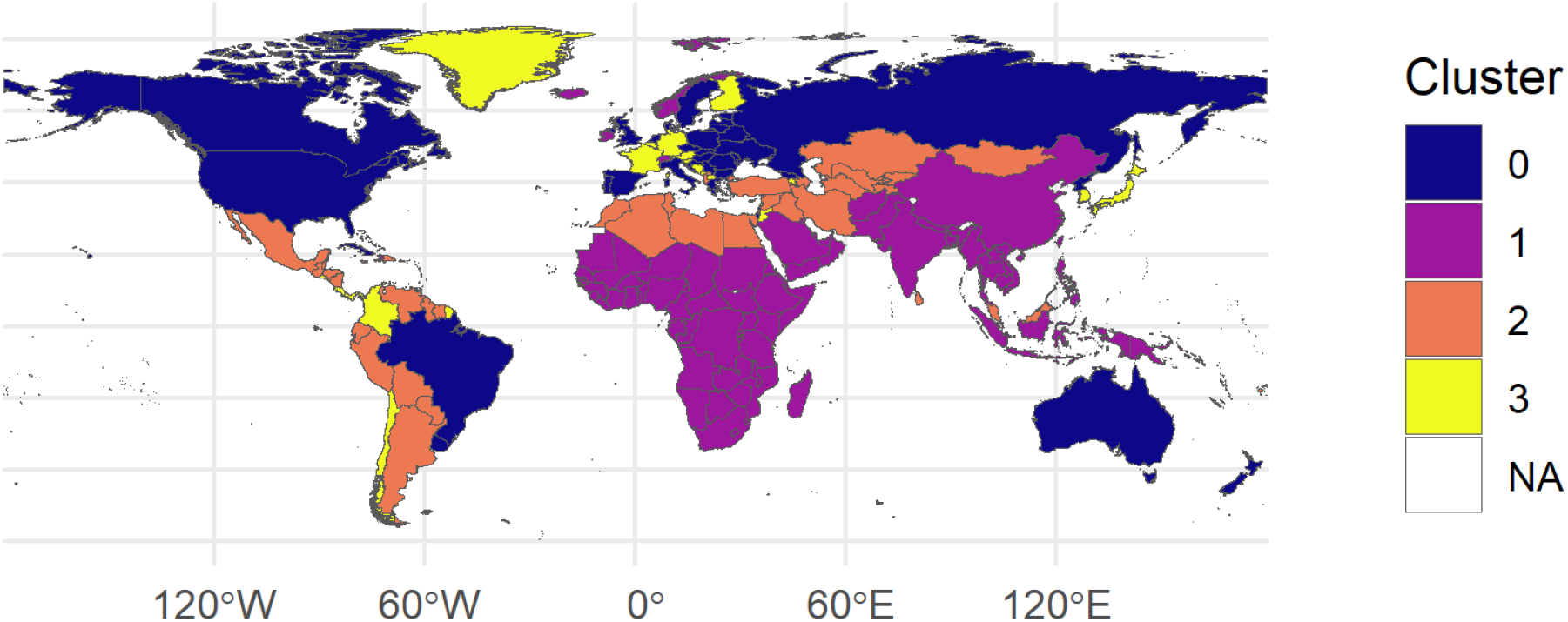
World map with cluster subdivision as described in Afuleni, Gada et al. (2025).

**Table A1.** **(a)** Lookup table of cluster-year combinations by estimate ID for A(H5N1). **(b)** Cluster memberships for the countries appearing in the WHO A(H5N1) dataset.

| ID | Cluster | Year |
| --- | --- | --- |
| 1 | Cluster0 | 2013 |
| 2 | Cluster0 | 2021 |
| 3 | Cluster0 | 2022 |
| 4 | Cluster0 | 2023 |
| 5 | Cluster0 | 2024 |
| 6 | Cluster1 | 2003 |
| 7 | Cluster1 | 2004 |
| 8 | Cluster1 | 2005 |
| 9 | Cluster1 | 2006 |
| 10 | Cluster1 | 2007 |
| 11 | Cluster1 | 2008 |
| 12 | Cluster1 | 2009 |
| 13 | Cluster1 | 2010 |
| 14 | Cluster1 | 2011 |
| 15 | Cluster1 | 2012 |
| 16 | Cluster1 | 2013 |
| 17 | Cluster1 | 2014 |
| 18 | Cluster1 | 2015 |
| 19 | Cluster1 | 2017 |
| 20 | Cluster1 | 2019 |
| 21 | Cluster1 | 2020 |
| 22 | Cluster1 | 2021 |
| 23 | Cluster1 | 2022 |
| 24 | Cluster1 | 2023 |
| 25 | Cluster1 | 2024 |
| 26 | Cluster2 | 2006 |
| 27 | Cluster2 | 2007 |
| 28 | Cluster2 | 2008 |
| 29 | Cluster2 | 2009 |
| 30 | Cluster2 | 2010 |
| 31 | Cluster2 | 2011 |
| 32 | Cluster2 | 2012 |
| 33 | Cluster2 | 2013 |
| 34 | Cluster2 | 2014 |
| 35 | Cluster2 | 2015 |
| 36 | Cluster2 | 2016 |
| 37 | Cluster2 | 2017 |
| 38 | Cluster2 | 2022 |
| 39 | Cluster3 | 2023 |

b)
| Cluster 0 | Cluster 1 | Cluster 2 | Cluster 3 |
| --- | --- | --- | --- |
| Australia | Bangladesh | Azerbaijan | Chile |
| Canada | Cambodia | Ecuador |  |
| Spain | Djibouti | Egypt |  |
| United Kingdom | India | Iraq |  |
| United States of America | Indonesia | Turkey |  |
|  | Lao PDR |  |  |
|  | Myanmar |  |  |
|  | Nepal |  |  |
|  | Nigeria |  |  |
|  | Pakistan |  |  |
|  | Thailand |  |  |
|  | Vietnam |  |  |

**Table A2.**
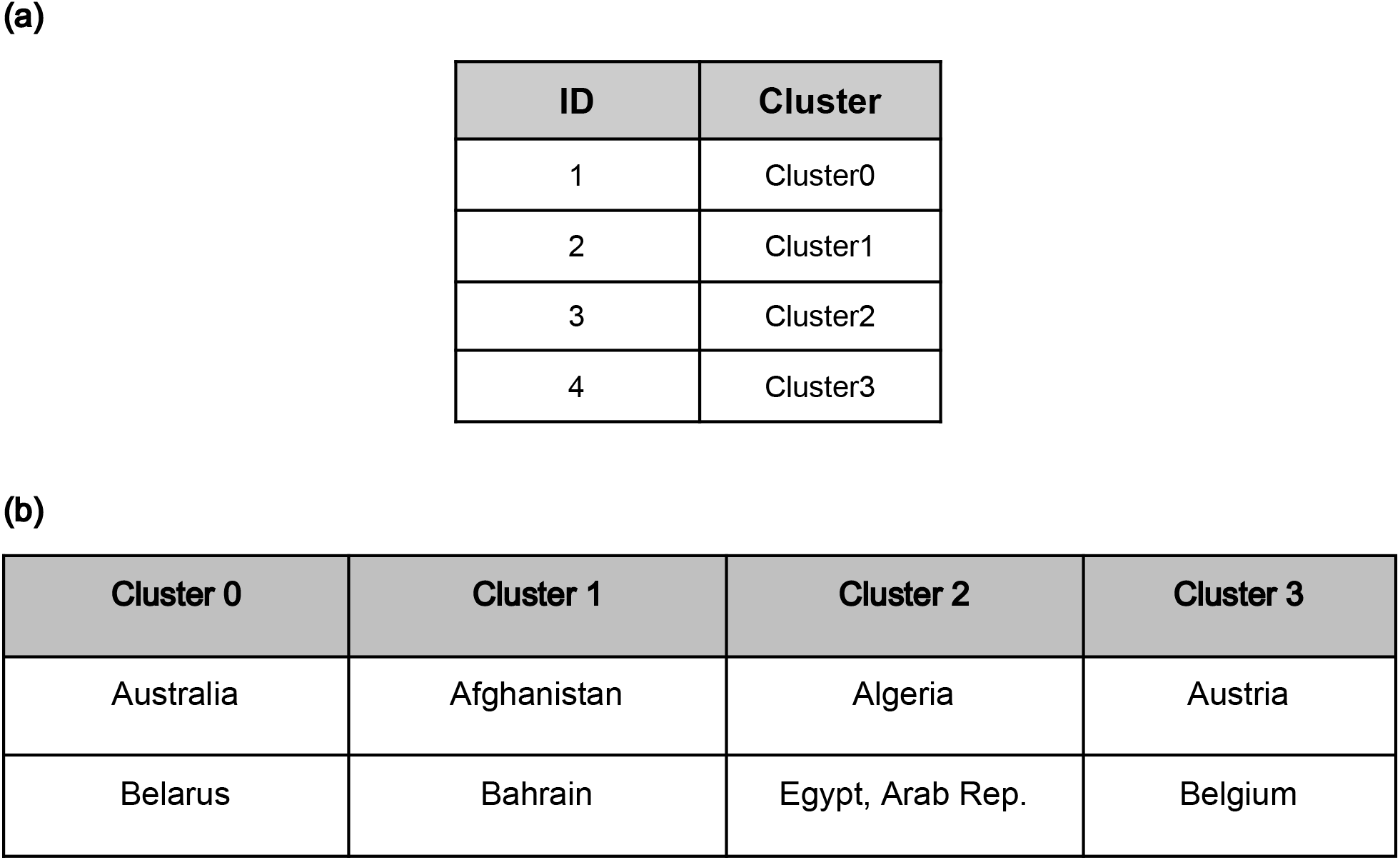
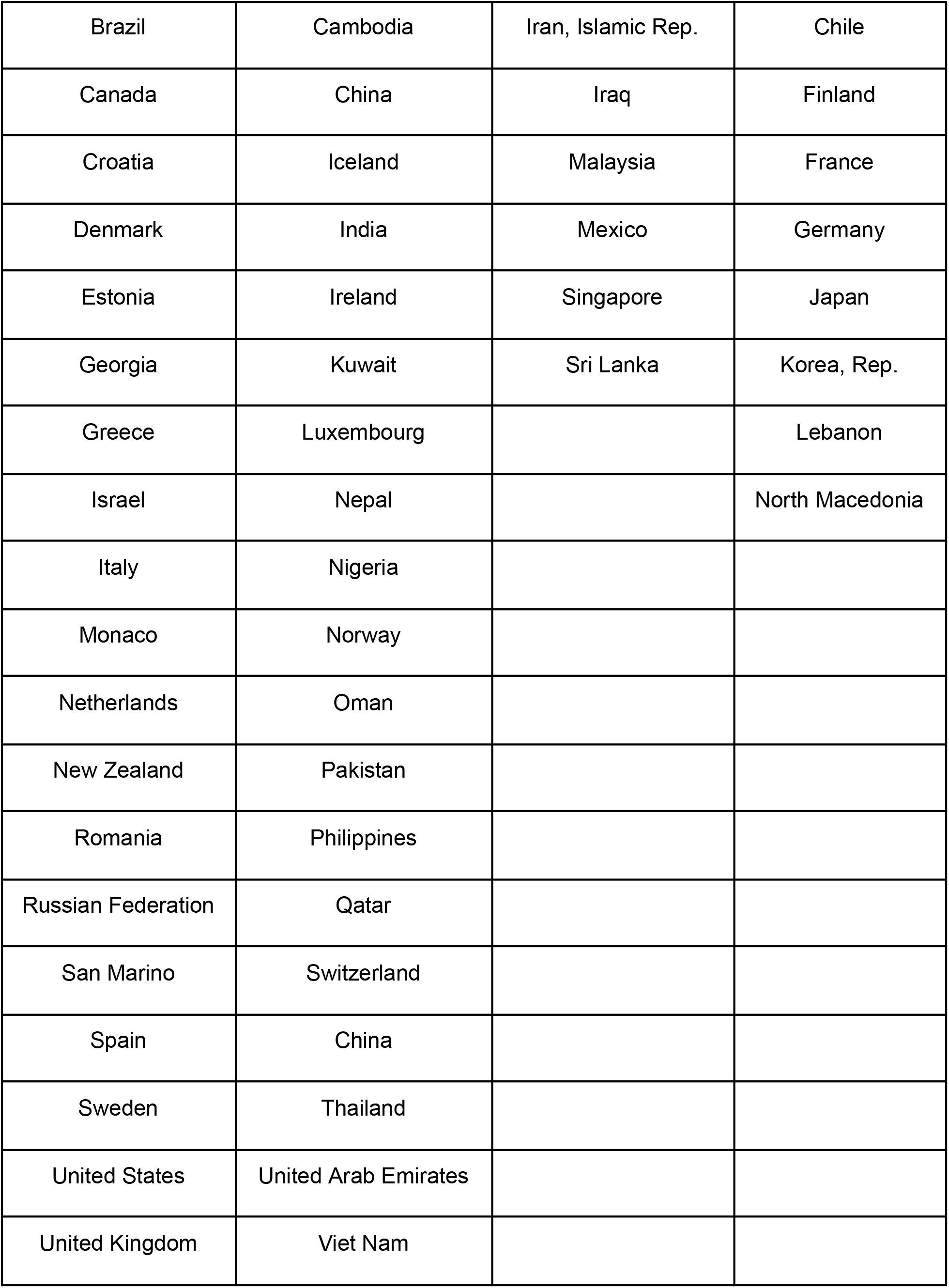
**(a)** Lookup table of cluster combinations by estimate ID for COVID-19. **(b)** Cluster memberships for the countries appearing in the COVID-19 dataset.

| ID | Cluster |
| --- | --- |
| 1 | Cluster0 |
| 2 | Cluster1 |
| 3 | Cluster2 |
| 4 | Cluster3 |

**(b)**
| Cluster 0 | Cluster 1 | Cluster 2 | Cluster 3 |
| --- | --- | --- | --- |
| Australia | Afghanistan | Algeria | Austria |
| Belarus | Bahrain | Egypt, Arab Rep. | Belgium |
| Brazil | Cambodia | Iran, Islamic Rep. | Chile |
| Canada | China | Iraq | Finland |
| Croatia | Iceland | Malaysia | France |
| Denmark | India | Mexico | Germany |
| Estonia | Ireland | Singapore | Japan |
| Georgia | Kuwait | Sri Lanka | Korea, Rep. |
| Greece | Luxembourg |  | Lebanon |
| Israel | Nepal |  | North Macedonia |
| Italy | Nigeria |  |  |
| Monaco | Norway |  |  |
| Netherlands | Oman |  |  |
| New Zealand | Pakistan |  |  |
| Romania | Philippines |  |  |
| Russian Federation | Qatar |  |  |
| San Marino | Switzerland |  |  |
| Spain | China |  |  |
| Sweden | Thailand |  |  |
| United States | United Arab Emirates |  |  |
| United Kingdom | Viet Nam |  |  |

## Notes

### Competing Interest Statement

The authors have declared no competing interest.

### Author Declarations

The source data for both cases and deaths were openly available on the World Health Organisation's website, John Hopkins University's COVID-19 GitHub repository, and the World Bank Database website.

