## Supplementary Diagnostic Plots for "Robustly Quantifying Uncertainty in International Avian Influenza A(H5N1) Infection Fatality Ratios": Supplementary Material_Rendered Plots.html


### Supplementary Material¶

This notebook is made available to complement the paper "Robustly Quantifying Uncertainty in International A(H5N1) Infection Fatality Ratios" by Gada et al. (2025). The code is an extract from the scripts available on GitHub at XXXXX, but we release this notebook to make it easier for readers to inspect the rendered diagnostic plots from the models without the need to rerun the analysis.

In [1]:

```
# Load required libraries
library(tidyverse)
library(cowplot)
library(posterior)
library(bayesplot)
library(rstan) # only for the purpose of this notebook for plotting
library(cmdstanr)
library(priorsense)
library(coda)
```

```
── Attaching core tidyverse packages ──────────────────────────────────────────────────────────────── tidyverse 2.0.0 ──
✔ dplyr     1.1.4     ✔ readr     2.1.5
✔ forcats   1.0.0     ✔ stringr   1.5.1
✔ ggplot2   4.0.1     ✔ tibble    3.3.0
✔ lubridate 1.9.4     ✔ tidyr     1.3.1
✔ purrr     1.0.2     
── Conflicts ────────────────────────────────────────────────────────────────────────────────── tidyverse_conflicts() ──
✖ dplyr::filter() masks stats::filter()
✖ dplyr::lag()    masks stats::lag()
ℹ Use the conflicted package (<http://conflicted.r-lib.org/>) to force all conflicts to become errors

Attaching package: 'cowplot'


The following object is masked from 'package:lubridate':

    stamp


This is posterior version 1.6.1.9000


Attaching package: 'posterior'


The following objects are masked from 'package:stats':

    mad, sd, var


The following objects are masked from 'package:base':

    %in%, match


This is bayesplot version 1.12.0

- Online documentation and vignettes at mc-stan.org/bayesplot

- bayesplot theme set to bayesplot::theme_default()

   * Does _not_ affect other ggplot2 plots

   * See ?bayesplot_theme_set for details on theme setting


Attaching package: 'bayesplot'


The following object is masked from 'package:posterior':

    rhat


Loading required package: StanHeaders

code for methods in class "Rcpp_model_base" was not checked for suspicious field assignments (recommended package 'codetools' not available?)

code for methods in class "Rcpp_model_base" was not checked for suspicious field assignments (recommended package 'codetools' not available?)

code for methods in class "Rcpp_stan_fit" was not checked for suspicious field assignments (recommended package 'codetools' not available?)

code for methods in class "Rcpp_stan_fit" was not checked for suspicious field assignments (recommended package 'codetools' not available?)


rstan version 2.32.6 (Stan version 2.32.2)


For execution on a local, multicore CPU with excess RAM we recommend calling
options(mc.cores = parallel::detectCores()).
To avoid recompilation of unchanged Stan programs, we recommend calling
rstan_options(auto_write = TRUE)
For within-chain threading using `reduce_sum()` or `map_rect()` Stan functions,
change `threads_per_chain` option:
rstan_options(threads_per_chain = 1)


Do not specify '-march=native' in 'LOCAL_CPPFLAGS' or a Makevars file


Attaching package: 'rstan'


The following objects are masked from 'package:posterior':

    ess_bulk, ess_tail


The following object is masked from 'package:tidyr':

    extract


This is cmdstanr version 0.9.0

- CmdStanR documentation and vignettes: mc-stan.org/cmdstanr

- CmdStan path: C:/Users/Leonardo.Gada/AppData/Local/miniforge3/envs/h5n1-modelling-env/Library/bin/cmdstan

- CmdStan version: 2.37.0


A newer version of CmdStan is available. See ?install_cmdstan() to install it.
To disable this check set option or environment variable cmdstanr_no_ver_check=TRUE.


Attaching package: 'coda'


The following object is masked from 'package:rstan':

    traceplot
```

#### A(H5N1) model results and diagnostics¶

Please refer to the script "H5N1 WHO Data CmdStanR v1.1.R" for the original code, including the data loading, cleaning and model running.

In [2]:

```
# Read in the outputs of the script "H5N1 WHO Data CmdStan v1.1.R"
# You should have run the code up to at least line 132 (just before the section "Routine Checks")

load("C:/Users/Leonardo.Gada/OneDrive - UK Health Security Agency/Documents/Modelling Projects/H5N1 CFR IFR/FinalGraphs/H5N1EnvironmentForPlots.RData")
```

In [9]:

```
# Traceplots to examine chain mixing and convergence
mcmc_trace(h5n1_model_draws, pars = names(h5n1_model_draws)[41:79], facet_args = list(ncol = 6)) + 
  theme(axis.text = element_text(size = 4)) +
  xlab("Post-warmup iteration") # check convergence diagnostics (IFR)
mcmc_trace(h5n1_model_draws, pars = names(h5n1_model_draws)[2:40], facet_args = list(ncol = 6)) + 
  theme(axis.text = element_text(size = 4)) + 
  xlab("Post-warmup iteration") # check convergence diagnostics (InfectionProb)
mcmc_trace(h5n1_model_draws, pars = names(h5n1_model_draws)[c(80,81,84:93)], facet_args = list(ncol = 4)) + 
  theme(axis.text = element_text(size = 4)) + 
  xlab("Post-warmup iteration") # check convergence diagnostics (alphas and betas std)
mcmc_trace(h5n1_model_draws, pars = names(h5n1_model_draws)[c(94:115)], facet_args = list(ncol = 4)) + 
  theme(axis.text = element_text(size = 4)) + 
  xlab("Post-warmup iteration") # check convergence diagnostics (gammas 1)
mcmc_trace(h5n1_model_draws, pars = names(h5n1_model_draws)[c(116:137)], facet_args = list(ncol = 4)) + 
  theme(axis.text = element_text(size = 4)) + 
  xlab("Post-warmup iteration") # check convergence diagnostics (gammas 2)
mcmc_trace(h5n1_model_draws, pars = names(h5n1_model_draws)[c(138:176)], facet_args = list(ncol = 6)) + 
  theme(axis.text = element_text(size = 4)) +  
  xlab("Post-warmup iteration") # check convergence diagnostics (infections)
mcmc_trace(h5n1_model_draws, pars = names(h5n1_model_draws)[c(177:215,82)], facet_args = list(ncol = 6)) + 
  theme(axis.text = element_text(size = 4)) + 
  xlab("Post-warmup iteration") # check convergence diagnostics (mu cases and phi cases)
mcmc_trace(h5n1_model_draws, pars = names(h5n1_model_draws)[c(216:254,83)], facet_args = list(ncol = 6)) + 
  theme(axis.text = element_text(size = 4)) + 
  xlab("Post-warmup iteration") # check convergence diagnostics (mu deaths and phi deaths)
mcmc_trace(h5n1_model_draws, pars = names(h5n1_model_draws)[c(255:293)], facet_args = list(ncol = 6)) + 
  theme(axis.text = element_text(size = 4)) +  
  xlab("Post-warmup iteration") # check convergence diagnostics (reporting prob cases)
mcmc_trace(h5n1_model_draws, pars = names(h5n1_model_draws)[c(294:332)], facet_args = list(ncol = 6)) + 
  theme(axis.text = element_text(size = 4)) + 
  xlab("Post-warmup iteration") # check convergence diagnostics (reporting prob deaths)
mcmc_trace(h5n1_model_draws, pars = names(h5n1_model_draws)[c(333:371)], facet_args = list(ncol = 6)) + 
  theme(axis.text = element_text(size = 4)) +  
  xlab("Post-warmup iteration") # check convergence diagnostics (true deaths)
mcmc_trace(h5n1_model_draws, pars = names(h5n1_model_draws)[c(372:421)], facet_args = list(ncol = 6)) + 
  theme(axis.text = element_text(size = 4)) + 
  xlab("Post-warmup iteration") # check convergence diagnostics (beta and gammas cases and deaths)
```

In [10]:

```
# Autocorrelation plots
mcmc_acf(h5n1_model_draws, pars = names(h5n1_model_draws)[41:51], lags = 10) # autocorrelation true ifr
mcmc_acf(h5n1_model_draws, pars = names(h5n1_model_draws)[52:65], lags = 10) # autocorrelation true ifr
mcmc_acf(h5n1_model_draws, pars = names(h5n1_model_draws)[66:79], lags = 10) # autocorrelation true ifr
mcmc_acf(h5n1_model_draws, pars = names(h5n1_model_draws)[2:14], lags = 10)  # autocorrelation infection prob
mcmc_acf(h5n1_model_draws, pars = names(h5n1_model_draws)[15:28], lags = 10) # autocorrelation infection prob
mcmc_acf(h5n1_model_draws, pars = names(h5n1_model_draws)[29:40], lags = 10) # autocorrelation infection prob
mcmc_acf(h5n1_model_draws, pars = names(h5n1_model_draws)[c(80,81,84:93)], lags = 10)  # autocorrelation alphas and betas std
mcmc_acf(h5n1_model_draws, pars = names(h5n1_model_draws)[94:104], lags = 10) # autocorrelation gammas
mcmc_acf(h5n1_model_draws, pars = names(h5n1_model_draws)[105:115], lags = 10) # autocorrelation gammas
mcmc_acf(h5n1_model_draws, pars = names(h5n1_model_draws)[116:126], lags = 10) # autocorrelation gammas
mcmc_acf(h5n1_model_draws, pars = names(h5n1_model_draws)[127:137], lags = 10) # autocorrelation gammas
mcmc_acf(h5n1_model_draws, pars = names(h5n1_model_draws)[138:149], lags = 10) # autocorrelation infections
mcmc_acf(h5n1_model_draws, pars = names(h5n1_model_draws)[150:162], lags = 10) # autocorrelation infections
mcmc_acf(h5n1_model_draws, pars = names(h5n1_model_draws)[163:176], lags = 10) # autocorrelation infections
mcmc_acf(h5n1_model_draws, pars = names(h5n1_model_draws)[c(82,177:189)], lags = 10) # autocorrelation mu and phi cases
mcmc_acf(h5n1_model_draws, pars = names(h5n1_model_draws)[190:202], lags = 10) # autocorrelation mu and phi cases
mcmc_acf(h5n1_model_draws, pars = names(h5n1_model_draws)[203:215], lags = 10) # autocorrelation mu and phi cases
mcmc_acf(h5n1_model_draws, pars = names(h5n1_model_draws)[c(83,216:226)], lags = 10) # autocorrelation mu deaths and phi deaths
mcmc_acf(h5n1_model_draws, pars = names(h5n1_model_draws)[227:240], lags = 10) # autocorrelation mu deaths and phi deaths
mcmc_acf(h5n1_model_draws, pars = names(h5n1_model_draws)[241:254], lags = 10) # autocorrelation mu deaths and phi deaths
mcmc_acf(h5n1_model_draws, pars = names(h5n1_model_draws)[255:268], lags = 10) # autocorrelation reporting prob cases
mcmc_acf(h5n1_model_draws, pars = names(h5n1_model_draws)[269:282], lags = 10) # autocorrelation reporting prob cases
mcmc_acf(h5n1_model_draws, pars = names(h5n1_model_draws)[283:293], lags = 10) # autocorrelation reporting prob cases
mcmc_acf(h5n1_model_draws, pars = names(h5n1_model_draws)[294:307], lags = 10) # autocorrelation reporting prob deaths
mcmc_acf(h5n1_model_draws, pars = names(h5n1_model_draws)[308:320], lags = 10) # autocorrelation reporting prob deaths
mcmc_acf(h5n1_model_draws, pars = names(h5n1_model_draws)[321:332], lags = 10) # autocorrelation reporting prob deaths
mcmc_acf(h5n1_model_draws, pars = names(h5n1_model_draws)[333:346], lags = 10) # autocorrelation true deaths
mcmc_acf(h5n1_model_draws, pars = names(h5n1_model_draws)[347:359], lags = 10) # autocorrelation true deaths
mcmc_acf(h5n1_model_draws, pars = names(h5n1_model_draws)[360:371], lags = 10) # autocorrelation true deaths
mcmc_acf(h5n1_model_draws, pars = names(h5n1_model_draws)[372:385], lags = 10) # autocorrelation beta and gammas cases and deaths
mcmc_acf(h5n1_model_draws, pars = names(h5n1_model_draws)[386:400], lags = 10) # autocorrelation beta and gammas cases and deaths
mcmc_acf(h5n1_model_draws, pars = names(h5n1_model_draws)[401:421], lags = 10) # autocorrelation beta and gammas cases and deaths
```

In [11]:

```
# Check energy of the No U-Turn algorithm
color_scheme_set("blue")
mcmc_nuts_energy(nuts_params(h5n1_model_fit), bins = 50) # check energy, https://arxiv.org/pdf/1701.02434
```

In [12]:

```
# Geweke Plots
h5n1_iter_mcmc <- as_mcmc.list(h5n1_model_fit) # convert CmdStanMCMC to MCMC list obj
geweke.plot(h5n1_iter_mcmc, frac1 = 0.50, frac2 = 0.50, ask = TRUE)   # mostly behave well
```

```
Error in plot.window(...): need finite 'ylim' values
Traceback:

1. plot(ystart, gcd[, j, k], type = "p", xlab = "First iteration in segment", 
 .     ylab = "Z-score", pch = 4, ylim = c(-ylimit, ylimit), ...)
2. plot.default(ystart, gcd[, j, k], type = "p", xlab = "First iteration in segment", 
 .     ylab = "Z-score", pch = 4, ylim = c(-ylimit, ylimit), ...)
3. localWindow(xlim, ylim, log, asp, ...)
4. plot.window(...)
5. .handleSimpleError(function (cnd) 
 . {
 .     watcher$capture_plot_and_output()
 .     cnd <- sanitize_call(cnd)
 .     watcher$push(cnd)
 .     switch(on_error, continue = invokeRestart("eval_continue"), 
 .         stop = invokeRestart("eval_stop"), error = NULL)
 . }, "need finite 'ylim' values", base::quote(plot.window(...)))
```

In [4]:

```
h5n1_sens <-powerscale_sensitivity(h5n1_model_fit,
                                   div_measure = "cjs_dist",
                                   component = c("prior", "likelihood"),
                                   sensitivity_threshold = 0.05,
                                   moment_match = FALSE,
                                   k_threshold = 0.5,
                                   transform = NULL,
                                   prediction = NULL)


filter(h5n1_sens, diagnosis == "potential prior-data conflict")
```

A powerscaled\_sensitivity\_summary: 52 × 4

| variable | prior | likelihood | diagnosis |
| --- | --- | --- | --- |
| <chr> | <dbl> | <dbl> | <chr> |
| infection\_prob[1] | 0.05269553 | 0.05269549 | potential prior-data conflict |
| infection\_prob[2] | 0.07044404 | 0.07044411 | potential prior-data conflict |
| infection\_prob[3] | 0.07964570 | 0.07964592 | potential prior-data conflict |
| infection\_prob[4] | 0.07102388 | 0.07102384 | potential prior-data conflict |
| infection\_prob[5] | 0.06585120 | 0.06585114 | potential prior-data conflict |
| infection\_prob[39] | 0.07880842 | 0.07880839 | potential prior-data conflict |
| true\_ifr[1] | 0.07021184 | 0.07021186 | potential prior-data conflict |
| true\_ifr[2] | 0.05951114 | 0.05951111 | potential prior-data conflict |
| true\_ifr[3] | 0.07911509 | 0.07911506 | potential prior-data conflict |
| true\_ifr[4] | 0.08693717 | 0.08693716 | potential prior-data conflict |
| true\_ifr[5] | 0.11208187 | 0.11208197 | potential prior-data conflict |
| true\_ifr[6] | 0.06725562 | 0.06725552 | potential prior-data conflict |
| true\_ifr[7] | 0.05505212 | 0.05505208 | potential prior-data conflict |
| true\_ifr[9] | 0.05000513 | 0.05000497 | potential prior-data conflict |
| true\_ifr[10] | 0.05529331 | 0.05529328 | potential prior-data conflict |
| true\_ifr[11] | 0.05757145 | 0.05757140 | potential prior-data conflict |
| true\_ifr[12] | 0.05858202 | 0.05858187 | potential prior-data conflict |
| true\_ifr[13] | 0.05405850 | 0.05405848 | potential prior-data conflict |
| true\_ifr[14] | 0.06411229 | 0.06411218 | potential prior-data conflict |
| true\_ifr[15] | 0.05914423 | 0.05914419 | potential prior-data conflict |
| true\_ifr[17] | 0.05413394 | 0.05413396 | potential prior-data conflict |
| true\_ifr[18] | 0.05134856 | 0.05134857 | potential prior-data conflict |
| true\_ifr[19] | 0.07959479 | 0.07959476 | potential prior-data conflict |
| true\_ifr[20] | 0.07862670 | 0.07862667 | potential prior-data conflict |
| true\_ifr[21] | 0.06506530 | 0.06506529 | potential prior-data conflict |
| true\_ifr[22] | 0.08056091 | 0.08056089 | potential prior-data conflict |
| true\_ifr[23] | 0.06378372 | 0.06378364 | potential prior-data conflict |
| true\_ifr[24] | 0.06714227 | 0.06714211 | potential prior-data conflict |
| true\_ifr[28] | 0.05782057 | 0.05782061 | potential prior-data conflict |
| true\_ifr[32] | 0.06028317 | 0.06028321 | potential prior-data conflict |
| true\_ifr[33] | 0.07461991 | 0.07461996 | potential prior-data conflict |
| true\_ifr[36] | 0.05728702 | 0.05728696 | potential prior-data conflict |
| true\_ifr[37] | 0.05951859 | 0.05951870 | potential prior-data conflict |
| true\_ifr[38] | 0.06677992 | 0.06677993 | potential prior-data conflict |
| true\_ifr[39] | 0.06404840 | 0.06404839 | potential prior-data conflict |
| infections[1] | 0.05269554 | 0.05269550 | potential prior-data conflict |
| infections[2] | 0.07044403 | 0.07044410 | potential prior-data conflict |
| infections[3] | 0.07964572 | 0.07964594 | potential prior-data conflict |
| infections[4] | 0.07102386 | 0.07102381 | potential prior-data conflict |
| infections[5] | 0.06585119 | 0.06585114 | potential prior-data conflict |
| infections[39] | 0.07880838 | 0.07880836 | potential prior-data conflict |
| mu\_deaths[3] | 0.05567270 | 0.05567268 | potential prior-data conflict |
| mu\_deaths[4] | 0.05929640 | 0.05929648 | potential prior-data conflict |
| mu\_deaths[5] | 0.08768994 | 0.08769012 | potential prior-data conflict |
| true\_deaths[3] | 0.06085903 | 0.06085902 | potential prior-data conflict |
| true\_deaths[4] | 0.06183598 | 0.06183606 | potential prior-data conflict |
| true\_deaths[5] | 0.08939670 | 0.08939689 | potential prior-data conflict |
| cases\_rep[21] | 0.05700210 | 0.05700209 | potential prior-data conflict |
| cases\_rep[39] | 0.05338232 | 0.05338230 | potential prior-data conflict |
| deaths\_rep[3] | 0.05080378 | 0.05080384 | potential prior-data conflict |
| deaths\_rep[4] | 0.06142652 | 0.06142656 | potential prior-data conflict |
| deaths\_rep[5] | 0.08938447 | 0.08938461 | potential prior-data conflict |

In [12]:

```
# we plot the variables that display potential prior-data conflict to examine whether this is the case
# the replicates we are not interested in, we only want to look at the parameters
powerscale_plot_dens(h5n1_model_fit, 
                     variable = c("infection_prob[1]","infection_prob[2]","infection_prob[3]",
                                "infection_prob[4]", "infection_prob[5]","infection_prob[39]"))
powerscale_plot_dens(h5n1_model_fit, 
                     variable = c("infections[1]","infections[2]","infections[3]",
                                "infections[4]", "infections[5]","infections[39]"))
powerscale_plot_dens(h5n1_model_fit, variable = c("mu_deaths[3]","mu_deaths[4]","mu_deaths[5]"))
powerscale_plot_dens(h5n1_model_fit, variable = c("true_deaths[3]","true_deaths[4]","true_deaths[5]"))
```

In [6]:

```
# look at all the true IFR
powerscale_plot_dens(h5n1_model_fit, variable = h5n1_sens$variable[40:50]) # need to split because graphic output is too big
```

In [5]:

```
powerscale_plot_dens(h5n1_model_fit, variable = h5n1_sens$variable[51:60])
```

In [7]:

```
powerscale_plot_dens(h5n1_model_fit, variable = h5n1_sens$variable[61:70])
```

In [8]:

```
powerscale_plot_dens(h5n1_model_fit, variable = h5n1_sens$variable[71:80])
```

In [13]:

```
# Posterior predictive checks
options(warn=-1)
cases_rep <- h5n1_model_fit$draws("cases_rep", format = "matrix") # workaround CmdStan output Env
# note that each row is a sample (predicted set of obs for each line in the dataset)
ppc_dens_overlay(y =clustered_data$ClusterCases, yrep = cases_rep) + xlim(c(0,1.5e3))

deaths_rep <- h5n1_model_fit$draws("deaths_rep", format = "matrix") # workaround CmdStan output Env
ppc_dens_overlay(y = clustered_data$ClusterDeaths, yrep = deaths_rep) + xlim(c(0,100))
options(warn=0)
```

In [12]:

```
# Raw output of bayesplot package
# Intervals and histograms
param <- names(h5n1_model_draws) # check column indexes of parameters
mcmc_hist(h5n1_model_fit$draws("true_ifr"), bins = 50) +
 theme(axis.text = element_text(size = 5))
mcmc_intervals(h5n1_model_fit$draws(format = "array"), 
               pars = param[41:79],
               prob = 0.50,
               prob_outer = 0.95)
```

#### COVID-19 model results and diagnostics¶

Please refer to the script "COVID Data CmdStanR v1.1.R" for the original code, including the data loading, cleaning and model running.

In [2]:

```
# Read in the outputs of the script "COVID Data CmdStan v1.1.R"
# You should have run the code up to at least line 164 (just before the section "Routine Checks"

load("C:/Users/Leonardo.Gada/OneDrive - UK Health Security Agency/Documents/Modelling Projects/H5N1 CFR IFR/FinalGraphs/CovidEnvironmentForPlots.RData")
```

In [21]:

```
# Traceplots to examine chain mixing and convergence
color_scheme_set("mix-brightblue-gray") # set the colour palette to shades of blue and gray
mcmc_trace(covid_model_draws, pars = names(covid_model_draws)[c(2:5,24:27)], facet_args = list(ncol = 3)) + 
  theme(axis.text = element_text(size = 6)) +
  xlab("Post-warmup iteration") # check convergence diagnostics (Infections and InfProb)
mcmc_trace(covid_model_draws, pars = names(covid_model_draws)[6:9]) + 
  theme(axis.text = element_text(size = 6)) +
  xlab("Post-warmup iteration") # check convergence diagnostics (IFR)
mcmc_trace(covid_model_draws, pars = names(covid_model_draws)[c(10:11,14:23,48:55)], facet_args = list(ncol = 4)) +
  theme(axis.text = element_text(size = 4)) +
  xlab("Post-warmup iteration") # check convergence diagnostics (alphas and betas)
mcmc_trace(covid_model_draws, pars = names(covid_model_draws)[c(12,13,28:35)], facet_args = list(ncol = 4)) + 
  theme(axis.text = element_text(size = 4)) +
  xlab("Post-warmup iteration") # check convergence diagnostics (MUs and PHIs)
mcmc_trace(covid_model_draws, pars = names(covid_model_draws)[36:47], facet_args = list(ncol = 4)) + 
  theme(axis.text = element_text(size = 4)) +
  xlab("Post-warmup iteration") # check convergence diagnostics (ReportingProbs and true deaths)
```

In [24]:

```
# Autocorrelation plots
mcmc_acf(covid_model_draws, pars = names(covid_model_draws)[c(2:5,24:27)], lags = 10)
mcmc_acf(covid_model_draws, pars = names(covid_model_draws)[c(6:9)], lags = 10)
mcmc_acf(covid_model_draws, pars = names(covid_model_draws)[c(10:11,48:55)], lags = 10)
mcmc_acf(covid_model_draws, pars = names(covid_model_draws)[c(14:23)], lags = 10)
mcmc_acf(covid_model_draws, pars = names(covid_model_draws)[c(12,13,28:35)], lags = 10)
mcmc_acf(covid_model_draws, pars = names(covid_model_draws)[c(36:47)], lags = 10)
```

In [10]:

```
# Check energy of the No U-Turn algorithm
mcmc_nuts_energy(nuts_params(covid_model_fit), bins = 50)
```

In [8]:

```
# Geweke Plots - this geweke.plot function is well described in the CODA manual
covid_iter_mcmc <- as_mcmc.list(covid_model_fit) # convert CmdStanMCMC to MCMC list obj
geweke.plot(covid_iter_mcmc, frac1 = 0.50, frac2 = 0.50, ask = TRUE)   # mostly behave well
```

In [4]:

```
# Power scaling checks

covid_sens <-powerscale_sensitivity(covid_model_fit,
                          div_measure = "cjs_dist",
                          component = c("prior", "likelihood"),
                          sensitivity_threshold = 0.05,
                          moment_match = FALSE,
                          k_threshold = 0.5,
                          transform = NULL,
                          prediction = NULL)
```

In [5]:

```
covid_sens
```

A powerscaled\_sensitivity\_summary: 62 × 4

| variable | prior | likelihood | diagnosis |
| --- | --- | --- | --- |
| <chr> | <dbl> | <dbl> | <chr> |
| infection\_prob[1] | 0.0099982597 | 0.0099982562 | - |
| infection\_prob[2] | 0.0106553710 | 0.0106553712 | - |
| infection\_prob[3] | 0.0107702414 | 0.0107702703 | - |
| infection\_prob[4] | 0.0109612936 | 0.0109612986 | - |
| true\_ifr[1] | 0.0124844841 | 0.0124844894 | - |
| true\_ifr[2] | 0.0118715633 | 0.0118715703 | - |
| true\_ifr[3] | 0.0125273443 | 0.0125273111 | - |
| true\_ifr[4] | 0.0126498590 | 0.0126498744 | - |
| alpha\_cases | 0.0003919995 | 0.0003920256 | - |
| alpha\_deaths | 0.0003262762 | 0.0003263391 | - |
| phi\_cases | 0.0008904651 | 0.0008904553 | - |
| phi\_deaths | 0.0008061612 | 0.0008062323 | - |
| beta\_cases\_std[1] | 0.0007767530 | 0.0007767634 | - |
| beta\_cases\_std[2] | 0.0005140594 | 0.0005140646 | - |
| beta\_cases\_std[3] | 0.0008341043 | 0.0008340883 | - |
| beta\_cases\_std[4] | 0.0004559318 | 0.0004559135 | - |
| b\_sigma\_cases | 0.0010270846 | 0.0010271117 | - |
| beta\_deaths\_std[1] | 0.0001809123 | 0.0001809119 | - |
| beta\_deaths\_std[2] | 0.0005070725 | 0.0005070850 | - |
| beta\_deaths\_std[3] | 0.0002988492 | 0.0002988679 | - |
| beta\_deaths\_std[4] | 0.0002128409 | 0.0002128336 | - |
| b\_sigma\_deaths | 0.0015419326 | 0.0015417943 | - |
| infections[1] | 0.0099983022 | 0.0099982988 | - |
| infections[2] | 0.0106553230 | 0.0106553233 | - |
| infections[3] | 0.0107702038 | 0.0107702327 | - |
| infections[4] | 0.0109612895 | 0.0109612944 | - |
| mu\_cases[1] | 0.0007640088 | 0.0007639869 | - |
| mu\_cases[2] | 0.0015391678 | 0.0015391556 | - |
| mu\_cases[3] | 0.0012690342 | 0.0012690497 | - |
| mu\_cases[4] | 0.0011972738 | 0.0011972570 | - |
| ⋮ | ⋮ | ⋮ | ⋮ |
| mu\_deaths[3] | 0.0001999803 | 0.0001999896 | - |
| mu\_deaths[4] | 0.0003609526 | 0.0003609213 | - |
| p[1] | 0.0019072195 | 0.0019072467 | - |
| p[2] | 0.0012526773 | 0.0012526981 | - |
| p[3] | 0.0024224428 | 0.0024224320 | - |
| p[4] | 0.0017111062 | 0.0017110644 | - |
| q[1] | 0.0003515326 | 0.0003515474 | - |
| q[2] | 0.0002307053 | 0.0002307176 | - |
| q[3] | 0.0003018049 | 0.0003018394 | - |
| q[4] | 0.0003610260 | 0.0003610441 | - |
| true\_deaths[1] | 0.0031422131 | 0.0031422380 | - |
| true\_deaths[2] | 0.0032118205 | 0.0032118175 | - |
| true\_deaths[3] | 0.0033057960 | 0.0033058250 | - |
| true\_deaths[4] | 0.0031533900 | 0.0031534151 | - |
| beta\_cases[1] | 0.0009447306 | 0.0009447399 | - |
| beta\_cases[2] | 0.0009882487 | 0.0009882559 | - |
| beta\_cases[3] | 0.0011396832 | 0.0011396725 | - |
| beta\_cases[4] | 0.0009000540 | 0.0009000327 | - |
| beta\_deaths[1] | 0.0002698260 | 0.0002698476 | - |
| beta\_deaths[2] | 0.0002231883 | 0.0002232003 | - |
| beta\_deaths[3] | 0.0002159065 | 0.0002159088 | - |
| beta\_deaths[4] | 0.0002314767 | 0.0002314865 | - |
| cases\_rep[1] | 0.0014017001 | 0.0014017002 | - |
| cases\_rep[2] | 0.0013129567 | 0.0013129568 | - |
| cases\_rep[3] | 0.0017912337 | 0.0017913091 | - |
| cases\_rep[4] | 0.0015411469 | 0.0015411471 | - |
| deaths\_rep[1] | 0.0017895366 | 0.0017897485 | - |
| deaths\_rep[2] | 0.0018674250 | 0.0018673191 | - |
| deaths\_rep[3] | 0.0030514089 | 0.0030516275 | - |
| deaths\_rep[4] | 0.0031793072 | 0.0031790062 | - |

In [4]:

```
#powerscale_plot_dens(covid_model_fit, variable = "true_ifr", facet_rows = "variable")
powerscale_plot_dens(covid_model_fit, variable = "true_ifr")
powerscale_plot_dens(covid_model_fit, variable = "infection_prob")
powerscale_plot_dens(covid_model_fit, variable = "infections")
powerscale_plot_dens(covid_model_fit, variable = "mu_cases")
powerscale_plot_dens(covid_model_fit, variable = "mu_deaths")
powerscale_plot_dens(covid_model_fit, variable = "true_deaths")
```

In [3]:

```
# Posterior predictive checks
options(warn=-1)

cases_rep <- covid_model_fit$draws("cases_rep", format = "matrix") # workaround CmdStan output Env
ppc_dens_overlay(y = covid_stan_data$TotClustCases, yrep = cases_rep) + xlim(0,2e3) # limited x axis loads to see real obs

deaths_rep <- covid_model_fit$draws("deaths_rep", format = "matrix") # workaround CmdStan output Env
ppc_dens_overlay(y = covid_stan_data$TotClustDeaths, yrep = deaths_rep) + xlim(c(0,100))
options(warn=-1)
```

In [5]:

```
# Raw output of bayesplot package
# Intervals and histograms
mcmc_hist(covid_model_fit$draws("true_ifr"), bins = 50)
mcmc_intervals(covid_model_fit$draws(format = "array"), 
               pars = c("true_ifr[1]", "true_ifr[2]", "true_ifr[3]", "true_ifr[4]"),
               prob = 0.50,
               prob_outer = 0.95)
```
